# Promoting Smoking Cessation and Preventing Relapse to Tobacco Use following a smokefree mental health in-patient stay (SCEPTRE feasibility study): a multi-centre randomised controlled feasibility study protocol

**DOI:** 10.1101/2024.10.31.24316412

**Authors:** Petal Petersen Williams, Lisa Huddlestone, Emily Shoesmith, Samantha Brady, Alex Mitchell, Victoria Exley, Fraser Wiggins, Lesley Sinclair, Jodi Pervin, Michelle Horspool, Moira Leahy, Claire Paul, Lesley Colley, Lion Shahab, Jude Watson, Catherine Hewitt, Simon Hough, John Britton, Tim Coleman, Simon Gilbody, Steve Parrott, Paul Galdas, Gregor Russell, Peter Coventry, Elena Ratschen

## Abstract

**Introduction:** Thousands of patients with mental illness are admitted to acute adult mental health wards every year in England, where local guidance recommends that all mental health settings be entirely smokefree. Mental health trusts presently invest substantial effort and resources to implement smokefree policies and to deliver tobacco dependence treatment to patients. Providing adequate support can help smokers remain abstinent or quit smoking during their smokefree inpatient stay and beyond. At present, little is known about how best to support patients to prevent their return to pre-admission smoking behaviours after discharge from a smokefree mental health inpatient stay. We have developed an intervention which includes targeted resources to support smoking-related behaviour change in patients following discharge from a smoke-free mental health setting. The aim of this trial is to determine the feasibility of a large-scale clinical trial to test the effectiveness and cost-effectiveness of the SCEPTRE intervention, compared with usual care.

**Methods and Analysis:** This feasibility study will be an individually randomised, controlled trial in eight National Health Service (NHS) mental health trusts recruiting adults (≥18 years) admitted to an acute adult mental health inpatient setting who are tobacco smokers on admission, or at any point during their inpatient stay. Consenting participants will be randomised to receive a 12-week intervention consisting of components aimed at promoting or maintaining positive smoking-related behaviour change following discharge from a smoke-free mental health inpatient setting or usual care. Data will be collected at baseline, 3-months and a second timepoint between 4-6 months post-randomisation. With 64 participants (32 in each group) the trial will allow a participation rate of 15% and completion rate of 80% to be estimated within a 95% confidence interval of ±3% and ±10% respectively. The analysis will be descriptive and follow a prespecified plan.

**Ethics and Dissemination:** Ethics approval was obtained from the North West – Greater Manchester West Research Ethics Committee. We will share results widely through local, national and international academic, clinical and Patient and Public Involvement (PPI) networks. The results will be disseminated through conference presentations, peer-reviewed journals and will be published on the trial website: https://sceptreresearch.com/.

**Trial registration number:** ISRCTN77855199

**Strengths and limitations of this study:**

- The use of a theory and evidence-based intervention to support smoking cessation and prevent relapse to tobacco following a smokefree mental health inpatient stay, can be effective for impacting long term smoking behaviour.
- The objective of the SCEPTRE feasibility trial is to test recruitment and radomisation of participants and collection of proposed outcome data in planning for a large randomised controlled trial.
- Quantitative and qualitative methods will be used to determine if research and intervention processes are acceptable and feasible and inform the decision of progression to full trial.
- The outcomes to be assessed will be relevant to patients, carers, mental health professionals, and policymakers.
- The present study is limited to English speaking adults admitted to acute mental health inpatient wards.
- Usual care varies greatly within and between Trusts nationally, making comparisons between intervention and standard care heterogenous.

## INTRODUCTION

Tobacco smoking remains one of the leading preventable causes of death and disease in England and is responsible for an estimated 74,600 deaths annually [1, 2]. Although smoking prevalence in the UK has steadily declined over the last few decades, rates remain at least 50% higher for people with mental health conditions compared to those without [3]. With average smoking prevalence figures of 40%, people with mental illness are more than twice as likely to smoke compared to those without [4]. Even higher smoking rates of up to 70% are seen in subgroups, such as hospitalised patients with mental health conditions [4, 5]. This results in substantially increased risks of premature smoking-related morbidity and mortality in this population [4]. Up to 20 life years are lost largely due to diseases related to smoking and this is the biggest contributor to the health inequalities experienced by people with mental illness [6].

Although people with mental health conditions are able [7] and more likely to be motivated [8] to quit smoking than those without, mainstream stop-smoking services are not commonly accessed by this population [9, 10] and are decreasingly resourced to support the needs of smokers with mental illness for tailored support [11–13]. However, National Institute for Health and Care Excellence (NICE) [14] guidance recommends that all mental health settings be entirely smokefree without exemption, with no facilitated smoking breaks, and evidence-based tobacco dependence treatment for smoking cessation, harm reduction and support for temporary abstinence available to all patients who smoke.

For many people with mental illness, a smokefree inpatient stay constitutes a rare or first experience as an adult of sustained abstinence, near-abstinence, or substantial reduction of tobacco consumption [15]. Evidence suggests individuals can successfully remain abstinent during their smokefree inpatient stay when behavioural and/or pharmacological support is offered [15, 16]. However, while a smokefree stay may result in temporary abstinence from tobacco or cessation, the risk of relapse post-discharge is high [17] with many individuals returning to smoking within a few days [18]. A lack of support in the immediate post-discharge period and the resulting relapse or return to heavy prehospital smoking patterns renders efforts and resources during the inpatient episode inefficient, as positive smoking behaviour change achieved during the inpatient stay may be lost. Therefore, it is vital to provide support post-discharge to prevent relapse.

The overall aim of the Promoting Smoking Cessation and Preventing Relapse to Tobacco use following a smokefree mental health in-patient stay (SCEPTRE) study is to determine the feasibility and acceptability of delivering the multi-component SCEPTRE intervention in National Health Service (NHS) adult mental health services. A second objective is to describe the initial effect of this intervention on participants’ smoking-related outcomes. We will also carry out a process evaluation and obtain feedback from all stakeholders relating to the research participation process and the acceptability of the intervention and gain an understanding of the importance and potential impact of individual intervention components. Finally, we will estimate the cost of delivering the SCEPTRE intervention and the control condition and the feasibility of collecting health economic data.

## METHODS AND ANALYSIS

The SCEPTRE feasibility trial protocol is reported in accordance with the guidelines presented in the Standard Protocol Items: Recommendations for Interventional Trials (SPIRIT) checklist [19] and the Template Intervention Description and Replication guidelines [20]. See supplemental appendix 1 for SPIRIT checklist.

### Patient and public involvement (PPI)

The SCEPTRE PPI group is integral to the trial and consists of six current/recent smokers with lived experience of a mental health inpatient stay and their carers. Members have contributed to the design of the intervention components, participant study documents, intervention resources, and intervention delivery mechanisms and measures. The group meets quarterly to provide feedback on study progress and have been kept up to date with frequent trial updates. Additionally, the Programme Steering Committee (PSC) has an independent lay member representative. The results of the study will be made available to trial participants, PPI panel members, participating NHS trusts and the wider public through the trial website and other popular media.

### Trial design

An individually randomised, controlled feasibility trial will be conducted with embedded qualitative interviews and health economic component (see Consolidated Standards of Reporting Trials (CONSORT) diagram in Fig. 1).

### Study setting

Participants will be recruited from up to eight English NHS mental health Trusts. To be eligible to participate, trusts must be committed to, and working towards full implementation of smokefree policy in line with national guidance [14].

### Participants

To be eligible for inclusion in this study, participants must (1) be aged 18 years and older (no maximum age); (2) be admitted to an acute adult mental health inpatient ward; (3) if detained under the mental health act, be granted unescorted (Section 17) leave i.e. leave the ward without being supervised (4) have discharge planned to an address/accommodation within the trust’s catchment area or a pre-defined radius agreed with participating trusts; (5) be a tobacco smoker at time of admission or at any point after (as patients occasionally still commence smoking after admission) and express an interest in maintaining abstinence if smokefree at time of assessment. Or express an interest in positively changing their smoking behaviour following discharge (including reduction of cigarette consumption; using e-cigarettes); (6) be able to understand and communicate in English; (7) have access to a telephone or computer/alternative digital device to receive post-discharge support; and (8) be willing and able to provide informed consent. Patients currently being treated within a psychiatric intensive care unit (PICU) will not be recruited. Additionally, those deemed not suitable to being approached to participate in the study (at clinician discretion) and those discharged to any trust accommodation with a smokefree policy will be ineligible to participate.

### Participant recruitment and study procedures

#### Patient identification and screening for eligibility

Participating trusts will identify, from electronic patient records, all patients who have been recorded on or after admission as a current smoker. Trust-based researchers will also visit participating wards and work with ward staff and other healthcare professionals to identify potential participants meeting the inclusion criteria. Potentially eligible participants will be approached by a member of the Trust research team to ascertain their interest in the study and seek permission to provide further information about the research. Screened patients will be recorded, along with reasons for ineligibility, non-approach and non-consent, in the eligibility screening log.

Ward staff will be encouraged to actively promote the study, for example, during ward community meetings and interactions with patients (e.g., during pre-discharge assessments and discharge planning). Patients who are identified as potential participants but who are too unwell to participate will be frequently reviewed to allow participation in the study when their mental health allows.

#### Informed consent and data collection

Consent will be obtained in-person or remotely via telephone/video call. All participants will have received a copy of the PIS at least 24 hours prior to obtaining consent. The consent form will also contain optional statements in relation to consent to interview for the qualitative aspect of the study.

Once participant eligibility has been confirmed and consent obtained, participants will be asked to complete a baseline questionnaire. If required, consent procedures and collection of baseline data can be separated into two visits dependent on participant preference. Data will be collected either using bespoke case report forms (CRFs) completed electronically via the secure web-based outcome data collection interface ‘REDCap’, over the telephone with a SCEPTRE researcher, or collected on paper CRFs returned via free post envelopes to York Trials Unit (YTU). All reporting of data collection will be undertaken in line with the CONSORT statement [22]. The baseline assessment includes socio-demographic information as well as information on mental and physical health and smoking history and behaviour. Participants who self-report they have not smoked will be asked to provide an expired air carbon monoxide (CO) reading. We will ask participants for full contact details at baseline (including mobile phone number, email, and address), and any contact preferences.

After the baseline assessment is completed, a member of the trust’s research team will randomise participants to either the intervention or comparison group. All participants (intervention and control) will be followed up for the purposes of the study via self-completed questionnaires at 3 months and 4-6 months post-randomisation to accommodate a variable second follow-up period which allows us to extend the recruitment period without extending the duration of the study overall. During the follow-up, outcome measures to be collected include smoking-related measures and mental and physical health measures. While completing the follow-up questionnaires, participants who self-report that they have not smoked in the previous seven days will be invited to undertake a CO measurement to validate their abstinence from tobacco. Participants who decline the invitation will be asked to provide a reason for declining, and this will be recorded on the questionnaire.

At three months and the relevant follow up timepoints, a link to complete the relevant electronic questionnaire on REDCap will be sent to participants via email, with the option to send a paper copy to participants for postal completion or completion with a researcher over the phone or in person instead as preferred. If participants select self-completion without a researcher and report abstinence from tobacco use within the previous 7 days, then a researcher will contact the participant to arrange a suitable time to obtain a CO reading in person to validate self-report. If no response is received within one week, an automated reminder will be sent to the participant. If participants do not respond following the automated reminder, a member of the research team will contact the participant via their preferred contact preference to prompt completion. Participants will receive shopping vouchers for completing the two follow-up assessments.

#### Qualitative interviews

Participants from both the control and intervention arms will be invited to take part in a short semi-structured interview following the completion of the SCEPTRE trial to gain a more in-depth understanding of the acceptability and feasibility of the study procedures. Interviews, guided by a schedule of topics developed from the APEASE criteria will examine the intervention delivery process, facilitators, and barriers of delivering the intervention, as well as the obtaining feedback from intervention arm participants. This will allow for future refinement and improvement of the intervention and research processes involved. Between nine and 12 patient participants in total (6-8 participants from the intervention arm; 3-4 participants from the control arm) will be interviewed. Up to four informal caregivers that can include family and friends (but not exclusively) of participants in the intervention arm will be invited to participate in short semi-structured interviews at 3-month follow-up to obtain their views on the acceptability and feasibility of intervention delivery methods. Semi-structured interviews will also be conducted with the leading mental health worker delivering the intervention from each of the Trusts in the week following the completion of the final participant in the intervention. Interviews will explore their experience and perceptions of delivering the intervention and assess the fidelity of delivery. Up to six trust stakeholders will be invited to participate in online focus group discussions (FGDs) to explore the experiences and perceptions of trust-based stakeholders of supporting the conduct of the research.

### Randomisation and blinding

Randomisation will be 1:1 to either the intervention or comparison arm, stratified by recruiting site and using randomly permuted blocks of randomly varying sizes. The randomisation schedule will be generated by a statistician at YTU not involved in the recruitment of participants and will be implemented in the REDCap system to ensure allocation concealment. Following a baseline assessment, randomisation will be undertaken by a member of the Trust’s research team. Due to the nature of the SCEPTRE intervention, the trust-based researchers and participants will not be blinded to allocation. The trial statistician conducting the analyses will not be blinded. Following allocation to a study arm, the researcher will receive an email with the participant’s allocation attached. The outcome of the allocation will be communicated to the participant where possible in person but may also be communicated by text or telephone call. Those participants randomised to the intervention arm will be contacted by the interventionist to arrange a time to complete the pre-discharge assessment. Details of participants allocated to the control group will be provided to ward staff to allow for the delivery of the usual tobacco dependency support offered by the trust. All participants’ GPs will be informed of their allocation by letter. A copy of the GP letter will also be held in the Trust electronic notes, so Community Mental Health Teams (CMHTs) are aware of patients’ participation in the study.

### Intervention

#### Control arm: usual care

Participants randomised to the control arm will receive usual care. The local offer of smoking cessation support is variable across NHS mental health trusts. Usual care in some trusts may be comprehensive and include behavioural and pharmacological support during admission, whereas others may provide limited support (e.g., access to nicotine replacement therapy). Detailed information on what usual care entails on each inpatient ward will be collected. All participants will be provided with information of how to access Stop Smoking Services post-discharge.

#### Intervention arm: SCEPTRE intervention package

Participants assigned to the intervention group will receive a 12-week intervention consisting of components aimed at promoting or maintaining smoking-related behaviour change among patients following discharge from a smoke-free mental health inpatient setting. See Fig. 2 for the SCEPTRE intervention pathway. The intervention development process was guided by the Behaviour Change Wheel model [23] and theoretically underpinned by the Theoretical Domains Framework [24] and the Behavioural Change Technique taxonomy [25]. Intervention components were identified and further developed based on two systematic reviews and a Delphi-style consultation process with key stakeholders, including clinicians and patients [16, 21, 26]. To ensure fit within the context of mental health services, refinement of the draft intervention was undertaken in collaboration with clinicians and experts in the field of tobacco control, and members of the SCEPTRE PPI panel. A more detailed description of the intervention and its development is provided elsewhere [21]. The intervention will be delivered by trained mental health workers, named ‘My-Try Specialists’ (MTSs). The manualised intervention consists of several core and additional components outlined in Table 1.

**Table 1.**
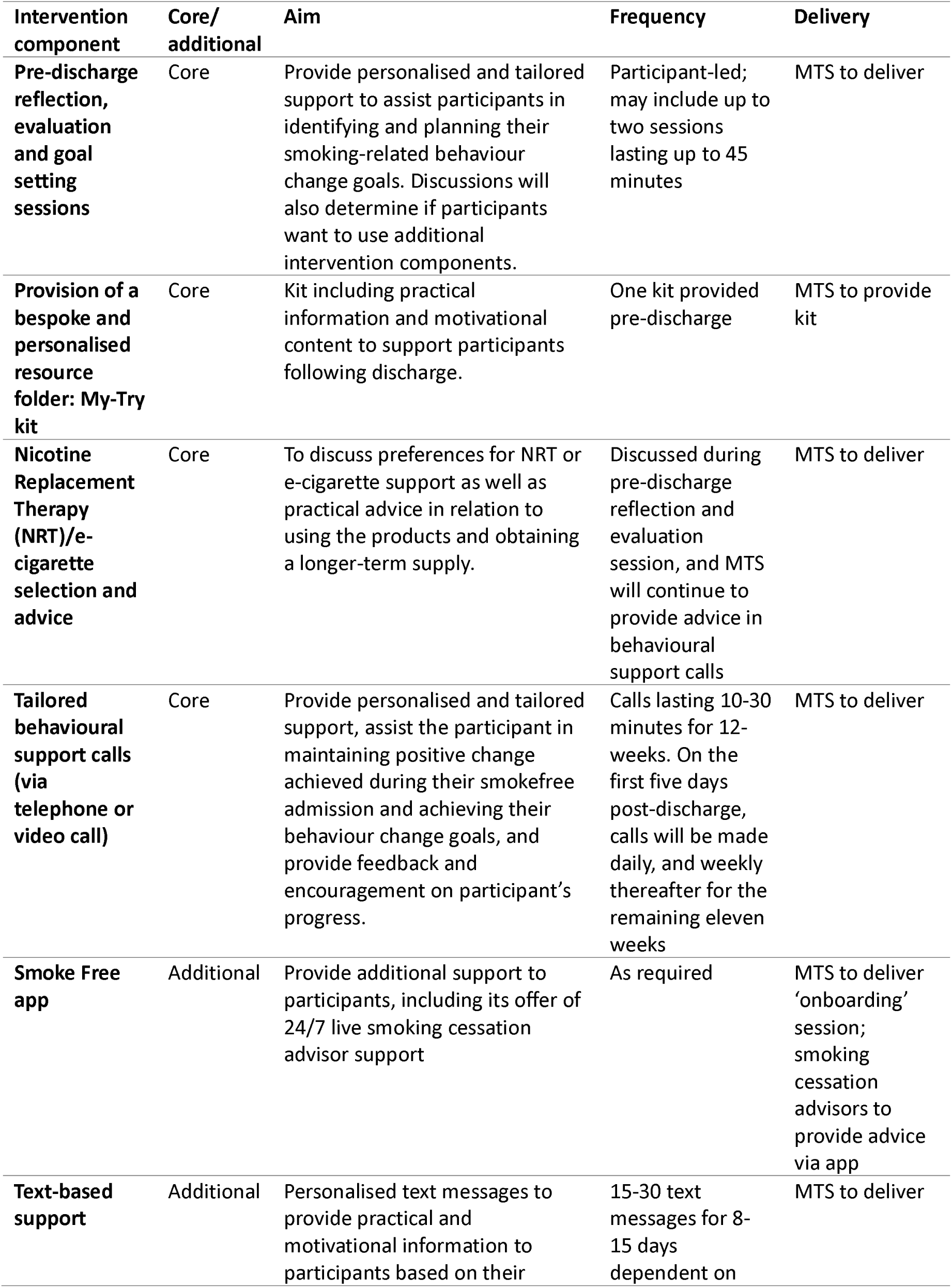
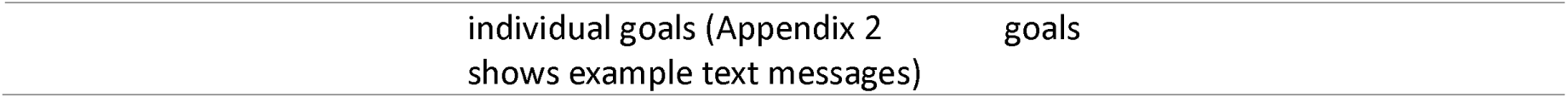
Summary of individual SCEPTRE intervention components.

### Interventionist training, supervision and competency

The MTS role aims to provide patients with tailored behavioural and social support and information to enable the continued change in smoking behaviours following discharge from a smoke-free mental health ward. To fulfil the MTS role, prospective interventionists are required to have knowledge of evidence-based methods in the treatment of tobacco dependency, including completion of the National Centre for Smoking Cessation Training (NCSCT) Practitioner Training module and the speciality course on smoking cessation and mental health. A bespoke training session designed to address the practicalities of smoking behaviour change in people with mental illness, and which covers e-cigarette use and interactions between tobacco smoke and certain antipsychotic medications (e.g., clozapine) in line with Royal College of Psychiatrists guidance, [27, 28] will be delivered.

MTSs will undertake self-guided online training supplemented with an in-person training session on the delivery of the manualised SCEPTRE intervention facilitated by members of the core SCEPTRE research team. Both clinical supervision, provided by the TRUST PI or nominated individual and procedural supervision, provided by members of the core SCEPTRE research team, on a fortnightly basis will be provided throughout the interventional period of the study. Fidelity of intervention delivery will be optimised through monthly supervision sessions and monitored using self-assessment checklists and logs for MTSs delivering the intervention. The log will allow MTSs to record all contacts with participants, and the research team will judge the degree to which the intervention as designed has been delivered in practice. Additional random fidelity checks will be carried out.

### Outcome measures and progression criteria

The schedule of data collection is shown in (Fig. 3) the SPIRIT Figure.

### Primary feasibility outcomes

The primary outcome of this feasibility trial will be to assess whether prespecified progression criteria are met to progress to the full randomised trial of the SCEPTRE intervention. Progression criteria are based on participant recruitment, retention, number of trusts enrolled and the feasibility of collecting carbon monoxide readings (Table 2).

**Table 2.**
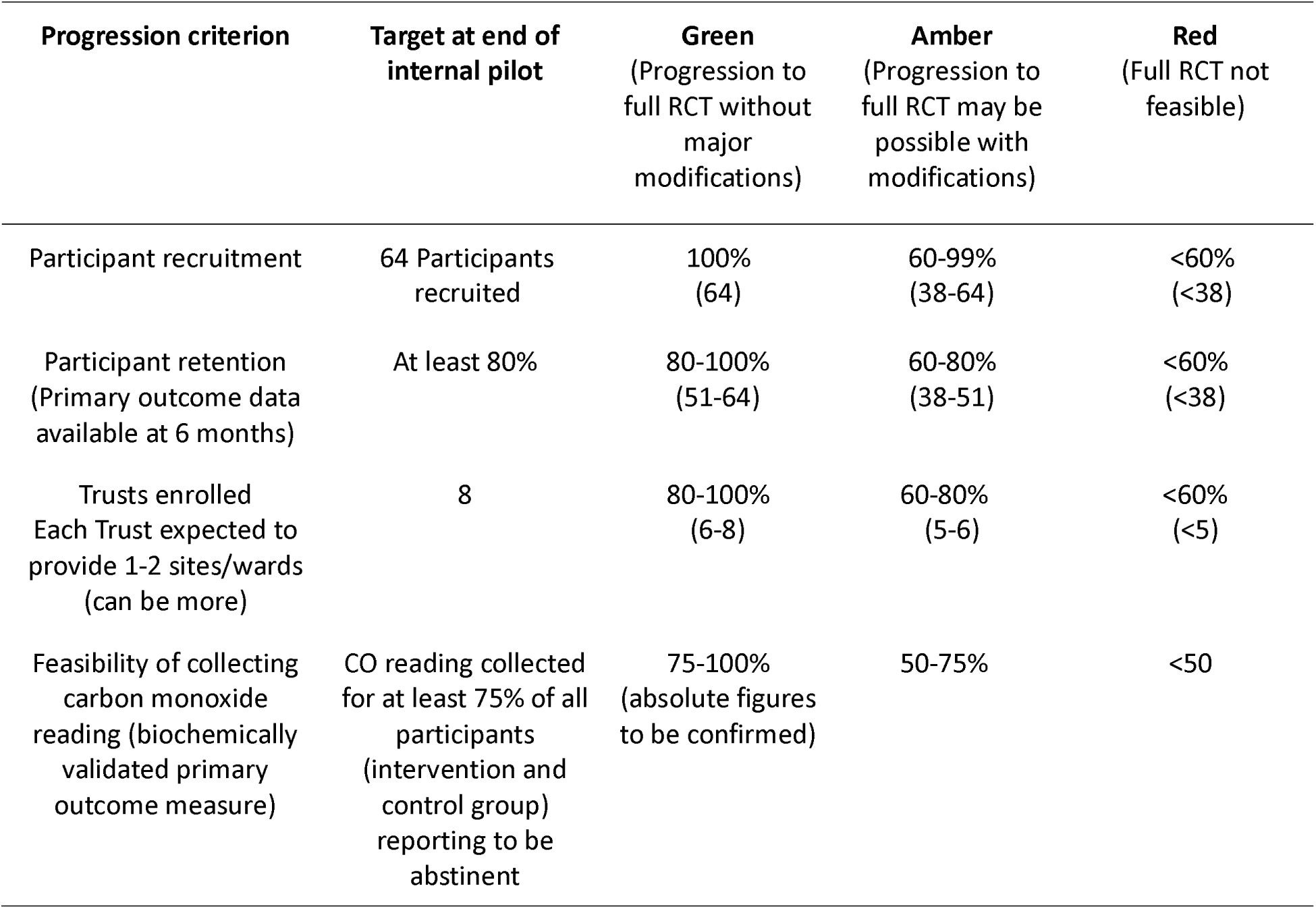
SCEPTRE feasibility study progression criteria.

### Proposed trial outcome measures

All participants will be asked to complete follow-up questionnaires with a member of the research team at 3– and either 4-, 5-, or 6-months post-randomisation (see Fig. 3).

#### Smoking-related measures

Smoking status; 7-day point prevalence abstinence validated by exhaled CO <10ppm, self-reported relapse following discharge from hospital, continuous abstinence, quit attempts, nicotine dependence as assessed by the Heaviness of Smoking Index (HSI) [30], strength of urges to smoke [31], level of motivation to quit as assessed by the Motivation to Stop Smoking Questionnaire [32] to assess how motivation changes over time and affects cessation outcomes, self-efficacy related to smoking cessation and use of e-cigarettes and NRT since discharge.

#### Mental and physical health measures

The nine-item Patient Health Questionnaire depression scale (PHQ-9) [33], the Generalised Anxiety Disorder 7-item scale (GAD-7) [34], and the quality-of-life scale (EQ-5D) [35], will be included. Frequency of use of health services (health economic data) since discharge will also be collected.

We will review the suitability and definitions of the two core smoking-related outcome measures, as per current debate in the field [36], as follows: for 7-day point prevalence abstinence, we will review and potentially adjust downward the CO threshold of <10 ppm for biochemical validation, depending on suitability of this approach for our patient population; for the multiple continuous abstinence outcome measures (1 – 6 months), we will review availability and congruency of data and define one consistent secondary outcome measure accordingly.

### Sample size considerations

The feasibility study will be conducted across approximately 12 acute adult mental health inpatient wards of the participating eight Trusts. Based on combined monthly discharge figures of ∼200 patients, of which we estimate conservatively that ∼140 will be eligible to participate, a smoking prevalence of ∼60%, and a conservative estimate of willingness of smokers to take part of 20%, we will aim to recruit a minimum of 64 patients, who will be individually randomised, at a 1:1 ratio, to the SCEPTRE intervention or the control arm. A trial of this size [37] will also allow a participation rate of 15% and completion rate of 80% to be estimated within a 95% confidence interval of ±3% and ±10% respectively [38].

### Additional qualitative and quantitative measures of feasibility and acceptability

In line with recommendations for the reporting of protocols of pilot and feasibility trials [29], a range of quantitative and qualitative data for all participants (intervention and control arm) determining the acceptability and feasibility of intervention delivery and research procedures will be collected.

#### Recruitment and retention

All participants screened will be recorded in the eligibility screening log, with reasons for ineligibility and approach outcomes (outlined above). Retention rates will be monitored throughout the duration of the study and the flow of participants will be detailed in a CONSORT diagram.

#### Compliance with the protocol

In addition to the interviews with MTSs (see above), compliance to the protocol and fidelity of intervention delivery will be optimised through monthly supervision sessions and monitored using self-assessment checklists and logs for MTSs delivering the intervention. The log will allow MTSs to record all contacts with participants, and the research team will judge the degree to which the intervention as designed has been delivered in practice. Any deviations from the protocol will be reported to YTU using a protocol deviation log.

#### Level of contamination and suitability of randomisaton approach

The suitability of the individual randomisation approach will be assessed by using qualitative and quantitative data of control group participants who quit or reduced smoking at 3– and 4, 5, or 6-months post-randomisation. Quantitative investigation will take place in the context of follow-up, to identify and specify smoking behaviour change. Qualitative data will be obtained via interviews (outlined above) to investigate reasons for smoking behaviour change and identify potential (unintentional) links with the SCEPTRE intervention. Based on our findings, we will estimate the presence/extent of contamination in the control group, and the need to change the unit of randomisation to clusters. If more than 30% of control participants appear to have changed aspects of their smoking behaviour due to factors directly related to our intervention, we will consider strategies to reduce contamination in the full RCT, including the possibility of designing the full RCT as a cluster trial, with wards as the unit of randomisation.

### Data management

Each site will hold data according to the General Data Protection Regulations (GDPR) and the Data Protection Act 2018. All data will be stored on a secure password-protected server and archived for 10 years after study completion. Patients will be assigned a unique trial number and this will be used on CRFs; patients will not be identified by their name in order to maintain confidentiality.

### Data analysis

A statistical analysis plan (SAP) giving details of the planned analyses will be drafted before data collection has been completed and reviewed by the Programme Management Group (PMG) and PSC. A brief description of the planned analyses is given below.

The statistical analysis will be carried out by a statistician based in YTU, using Stata v17 or later. The reporting of this trial will follow CONSORT guidelines for pilot and feasibility trials [22]. A flow diagram will be produced, depicting the flow of patients through the trial. The number of patients screened, eligible, consenting, and randomised will be summarised, with reasons for ineligibility and non-consent given where available. Baseline data will be summarised descriptively by randomised group, with no formal statistical comparisons being undertaken [39]. Continuous variables will be summarised using descriptive statistics, while categorical variables will be summarised using counts and percentages. Descriptive statistics will summarise participant outcomes by randomised group and timepoint, including the amount of missing data. Relevant measures of recruitment and retention will be calculated and compared against the pilot progression criteria. The recruitment rate will be estimated and presented alongside a corresponding 95% confidence interval. Measures of contamination and participant engagement with the multiple components of the intervention will be summarised descriptively.

Health economic analysis will be conducted as a preparatory step towards a comprehensive economic evaluation in the subsequent definitive RCT. Costs associated with the delivery of the SCEPTRE intervention and usual care will be estimated. All resources used in the provision of the intervention, such as the training for the MTSs, the staff time spent on delivering the intervention, and the NRT and e-cigarettes dispensed, will be recorded alongside the trial. Data on the resources used in usual care through self-completed questionnaires at 3– and either 4-, 5-, or 6-months post-randomisation will be collected. A micro-costing approach, collecting detailed information about health service use, will be applied to generate estimated intervention costs for both arms by multiplying the quantity of each identified resource by its corresponding unit cost. We will pilot the data collection tools used to gather health economics data. We will test the feasibility and acceptability of the health service use questionnaire and the outcome measure instrument (i.e., the EQ-5D-5L questionnaire). We will examine the completion rates, identify any challenges or barriers to completion, and make necessary modifications to the questionnaires for use in the full trial. The methods and findings will be summarised in a report that can be used to inform the design and conduct of the economic evaluation in the full trial stage.

Qualitative data analysis will be undertaken by group, e.g., participants, MTSs, friends/family, Trust stakeholders. Data will be analysed using NVivo software (QSR International Pty Ltd, Melbourne, Australia). First, an inductive approach using thematic analysis will be used to identify nodes and sub-themes in the data [40]. Transcripts will be coded line by line by two researchers, with preliminary code names assigned to the data items and iteratively developed. Preliminary codes will be checked and discussed with a third researcher, experienced in qualitative research and the behaviour change wheel. Following this, a deductive approach will be used to chart sub-themes to the APEASE (Acceptability, Practicability, Effectiveness, Affordability, Side-effects, and Equity) criteria. The data will be presented and discussed in a wider research team meeting to refine and confirm the final interpretation.

### Trial management

A Programme Management Group (PMG) has been established to oversee the day-to-day management (e.g., protocol and ethics approvals, set-up, recruitment, data collection, data management) of the study, and is chaired by the Chief Investigator (CI). Membership includes co-CIs, co-investigators, research staff on the project and PPI representation. The role of the PMG is to monitor all aspects of the conduct and progress of the trial, ensure that the protocol is adhered to and take appropriate action to safeguard participants and the quality of the trial itself. Throughout the project there will be regular videoconference contact supplemented by face-to-face meetings where required. Frequency of meetings will vary depending on the stage of the trial. The PMG, through the York Trials Unit (YTU), will provide feedback on trial progress to the Programme Steering Committee (PSC) established to provide overall independent oversight for SCEPTRE on behalf of the Sponsor and Project Funder and to ensure that the project is conducted to the rigorous standards set out in the Department of Health’s Research Governance Framework for Health and Social Care and the Guidelines for Good Clinical Practice (GCP). Furthermore, the PMG through the YTU, will provide feedback to the Data Monitoring and Ethics Committee (DMEC) established to monitor safety and efficacy data as well as quality and compliance data and ensure that the protocol is accurately followed, and the study is GCP compliant.

### Ethics

The protocol (Version 1.0_05.09.2023) for this study and the informed consent form (see online supplemental appendix 2) were approved by the North West – Greater Manchester West Research Ethics Committee (REC) (23/NW/0312). The Investigators will ensure that this study is conducted in full conformity with current regulations, the current revision of the Declaration of Helsinki, and with the principles of GCP. We will adhere to the UK Framework for Health and Social Care Research [22, 39]. The programme manager/CI will obtain approval for all amendments.

### Adverse events

All adverse events (AEs) occurring during the study observed by the investigator or reported by MTSs or participants, will be recorded on the SCEPTRE Adverse Event Form for return to YTU. All serious adverse events (SAEs) will be entered onto the SAE reporting form and sent via REDCap or encrypted email to YTU within 24 hours of the investigator becoming aware of the event. Once received, causality and expectedness will be confirmed by the CI or a medical co-applicant or PSC member not acting as a site Principal Investigator (PI). Any change of condition or other follow-up information should be sent as soon as it is available or at least within 24 hours of the information becoming available. Events will be followed up until the event has resolved or a final outcome has been reached. SAEs that are deemed to be unexpected and related to the trial will be notified to the REC and sponsor within 15 days by YTU. All such events will be reported to the PSC and DMEC at their next meetings. Protocols and SOPs have been developed to identify and manage risks of suicide and harm to participants.

### Dissemination

We will share results widely through local, national and international academic, clinical and PPI networks. The results will be disseminated through conference presentations, peer reviewed journals and will be published on the trial website: https://sceptreresearch.com/.

### Trial status

This protocol (V.1.0) was approved on 6^th^ November 2023. Recruitment to the SCEPTRE feasibility study opened on 23 January 2024 with the first participant randomised on 13^th^ February 2024. The trial is ongoing. Reporting of the trial is anticipated in the first quarter of 2025.

## DISCUSSION

In line with NICE recommendations, that mental health settings become entirely smokefree, and that mental health patients should have access to evidence-based stop smoking treatment [14], SCEPTRE aims to test ways to support mental health inpatients after discharge in maintaining or achieving abstinence from tobacco smoking. At present, no evidence-based strategies to help maintain or achieve a smokefree lifestyle and avoid relapse after discharge exist. As such, SCEPTRE feasibility study will inform the design of a future large-scale randomised controlled trial. This large-scale trial will aim to test the effectiveness and cost-effectiveness of a complex intervention to promote or maintain smoking-related behaviour change among patients following discharge from a smoke-free mental health inpatient setting. One limitation of this study is the potential for contamination in the control group. However, we will assess and estimate the presence/extent of this, and the need to change the unit of randomisation to clusters. Secondly, this is a multi-component intervention which makes it difficult to delineate impact very clearly. The process evaluation will aim to provide insights on the importance of individual intervention components. The study is focused on acute adult mental health departments making generalisation beyond this setting difficult. Usual care is heterogeneous and policy/practice landscapes are shifting rapidly. We will collect information on what usual care entails on each inpatient ward.

If effective and implemented in NHS mental health settings, the SCEPTRE intervention will directly benefit patients (and their families) and has the potential to save lives and NHS resources, and to decrease health inequalities that exists for people with mental health conditions.

## Supporting information

Fig 1 SCEPTRE CONSORT

Fig 2 SCEPTRE intervention pathway

Fig 3 SPIRIT

Supp 1 SPIRIT checklist

Supp 2 Consent Form

## Data Availability

All data produced in the present study are available upon reasonable request to the authors

## Acknowledgements

We wish to thank the SCEPTRE PPI Panel for their assistance in the development of patient/participant processes and materials. We also acknowledge the valuable contributions of the NCSCT for their assistance in the development and delivery of MTS training on smoking and mental health, and the Smoke Free App team for their contributions to the development of processes and materials to support participant engagement with the Smoke Free App.

## Authors’ Contributions

ER conceived the idea for the study. ER, LH, ES, LC, ML, and CP led the development of the intervention. VE is the Trial Co-ordinator and SB and LSinclair are the trial managers. AM and FW are the study statisticians. GR, SG, CH, TC, JW, SP, PC, MH, JB, ML, CP, PG, LShahab, LC and MH are co-applicants and SH is the independent PPI co-applicant. JB, PC, TC, PG, SG, CH, SP, LShahab, and JW provide academic expertise. PPW drafted the manuscript. All authors contributed to protocol writing and approved the final version.

## Funding statement

The SCEPTRE Feasibility Trial is funded by Programme Grants for Applied Research (NIHR200607). The sponsor is the Sheffield Health and Social Care NHS Foundation Trust. Catherine Hewitt, Simon Gilbody and Tim Coleman are NIHR Senior Investigators. The trial will be monitored and audited in accordance with the sponsor’s procedures. The sponsor will play no role in the study design; collection, management, analysis, and interpretation of data; writing of the report; and the decision to submit the report for publication. PPW, LH, ES, and ER are supported by the NIHR Yorkshire and Humber Applied Research Collaboration.

## Competing interest statement

LS has received honoraria for talks, an unrestricted research grant and travel expenses to attend meetings and workshops from Pfizer and an honorarium to sit on advisory panel from Johnson & Johnson, both pharmaceutical companies that make smoking cessation products. He has acted as paid reviewer for grant awarding bodies and as a paid consultant for health care companies. Other research has been funded by the Department of Health, UKRI, a community-interested company (National Centre for Smoking Cessation) and charitable sources (Cancer Research UK, Yorkshire Cancer Research). He has never received personal fees or research funding of any kind from alcohol, electronic cigarette or tobacco companies. All other authors declare no conflicts of interest.

## Notes

### Clinical Trial

ISRCTN77855199

## REFERENCES

1. Digital, N. Statistics on Smoking, England 2020. 2020 [cited 2023 08/08]; Available from: https://digital.nhs.uk/data-and-information/publications/statistical/statistics-on-smoking/statistics-on-smoking-england-2020/part-1-smoking-related-ill-health-and-mortality.

2. Digital, N., Statistics on Smoking, England – 2016 2016.

3. Richardson, S., A. McNeill, and L.S. Brose, Smoking and quitting behaviours by mental health conditions in Great Britain (1993–2014). Addictive behaviors, 2019. 90: p. 14–19.

4. Psychiatrists, R.C.o.P.R.C.o., Smoking and mental health. 2013: London.

5. Meltzer, H., et al., The prevalence of psychiatric morbidity among adults living in institutions. International Review of Psychiatry, 2003. 15(1-2): p. 129–133.

6. Cheeseman H H.H., The stolen years: The mental health and smoking action report 2016: London.

7. Peckham, E., et al., Smoking cessation in severe mental ill health: what works? An updated systematic review and meta-analysis. BMC psychiatry, 2017. 17(1): p. 1–18.

8. Brose, L.S., et al., Mental health, smoking, harm reduction and quit attempts – a population survey in England. BMC Public Health, 2020. 20(1): p. 1237.

9. McNally, L. and E. Ratschen, The delivery of stop smoking support to people with mental health conditions: a survey of NHS stop smoking services. BMC Health Services Research, 2010. 10: p. 1–5.

10. McNally, L., C. Todd, and E. Ratschen, The prevalence of mental health problems among users of NHS stop smoking services: effects of implementing a routine screening procedure. BMC health services research, 2011. 11(1): p. 1–4.

11. Knowles, S., et al., Making the journey with me: a qualitative study of experiences of a bespoke mental health smoking cessation intervention for service users with serious mental illness. BMC psychiatry, 2016. 16: p. 1–9.

12. Peckham, E., et al., Smoking Cessation Intervention for severe Mental Ill Health Trial (SCIMITAR): a pilot randomised control trial of the clinical effectiveness and cost-effectiveness of a bespoke smoking cessation service. Health Technology Assessment (Winchester, England), 2015. 19(25): p. 1–148, v.

13. Parker, C., A. McNeill, and E. Ratschen, Tailored tobacco dependence support for mental health patients: a model for inpatient and community services. Addiction, 2012. 107: p. 18–25.

14. NICE. Tobacco: preventing uptake, promoting quitting and treating dependence [NG209]. 2021; Available from: https://www.nice.org.uk/guidance/ng209.

15. Brose, L.S., E. Simonavicius, and A. McNeill, Maintaining abstinence from smoking after a period of enforced abstinence–systematic review, meta-analysis and analysis of behaviour change techniques with a focus on mental health. Psychological medicine, 2018. 48(4): p. 669–678.

16. Shoesmith, E., et al., Supporting smoking cessation and preventing relapse following a stay in a smoke-free setting: a meta-analysis and investigation of effective behaviour change techniques. Addiction, 2021. 116(11): p. 2978–2994.

17. Prochaska, J.J., et al., Return to smoking following a smoke-free psychiatric hospitalization. American Journal on Addictions, 2006. 15(1): p. 15–22.

18. Mussulman, L.M., et al., Rapid relapse to smoking following hospital discharge. Preventive medicine reports, 2019. 15: p. 100891.

19. Chan, A.W., et al., SPIRIT 2013 statement: defining standard protocol items for clinical trials. Ann Intern Med, 2013. 158(3): p. 200–7.

20. Alvarez, G., F. Cerritelli, and G. Urrutia, Using the template for intervention description and replication (TIDieR) as a tool for improving the design and reporting of manual therapy interventions. Man Ther, 2016. 24: p. 85–9.

21. Shoesmith, E., et al., Promoting and Maintaining Changes in Smoking Behavior for Patients Following Discharge from a Smoke-free Mental Health Inpatient Stay: Development of a Complex Intervention Using the Behavior Change Wheel. Nicotine and Tobacco Research, 2023. 25(4): p. 729–737.

22. Eldridge, S.M., et al., CONSORT 2010 statement: extension to randomised pilot and feasibility trials. bmj, 2016. 355.

23. Michie, S., L. Atkins, and R. West, The behaviour change wheel. A guide to designing interventions. 1st ed. Great Britain: Silverback Publishing, 2014. 1003: p. 1010.

24. Cane, J., D. O’Connor, and S. Michie, Validation of the theoretical domains framework for use in behaviour change and implementation research. Implementation science, 2012. 7: p. 1–17.

25. Michie, S., et al., The behavior change technique taxonomy (v1) of 93 hierarchically clustered techniques: building an international consensus for the reporting of behavior change interventions. Annals of behavioral medicine, 2013. 46(1): p. 81–95.

26. Huddlestone, L., et al., A systematic review of mental health professionals, patients, and carers’ perceived barriers and enablers to supporting smoking cessation in mental health settings. Nicotine and Tobacco Research, 2022. 24(7): p. 945–954.

27. Physicians, R.C.o. and R.C.o. Psychiatrists. Smoking and mental health 2013 [cited 2022 23 November]; Available from: https://www.rcplondon.ac.uk/projects/outputs/smoking-andmental-health

28. Royal College of Psychiatrists, The prescribing of varencline and vaping (eletronic cigarettes) to patients with severe mental illness. Position Statement (PS05/18). 2018.

29. Thabane, L. and G. Lancaster, A guide to the reporting of protocols of pilot and feasibility trials. Pilot and Feasibility Studies, 2019. 5(1): p. 37.

30. Borland, R., et al., The reliability and predictive validity of the Heaviness of Smoking Index and its two components: findings from the International Tobacco Control Four Country study. Nicotine & Tobacco Research, 2010. 12(suppl_1): p. S45–S50.

31. Fidler, J.A., L. Shahab, and R. West, Strength of urges to smoke as a measure of severity of cigarette dependence: comparison with the Fagerström Test for Nicotine Dependence and its components. Addiction, 2011. 106(3): p. 631–638.

32. (NCSCT), N.C.f.S.C.a.T. Motivation to Stop Smoking Available from: https://www.ncsct.co.uk/usr/pub/Motivation%20to%20stop%20smoking.pdf.

33. Kroenke, K., R.L. Spitzer, and J.B. Williams, The PHQ-9: validity of a brief depression severity measure. Journal of general internal medicine, 2001. 16(9): p. 606–613.

34. Spitzer, R.L., et al., A brief measure for assessing generalized anxiety disorder: the GAD-7. Archives of internal medicine, 2006. 166(10): p. 1092–1097.

35. Byford, S., The validity and responsiveness of the EQ-5D measure of health-related quality of life in an adolescent population with persistent major depression. Journal of mental health, 2013. 22(2): p. 101–110.

36. Benowitz, N.L., et al., Biochemical Verification of Tobacco Use and Abstinence: 2019 Update. Nicotine Tob Res, 2020. 22(7): p. 1086–1097.

37. Metse, A.P., et al., Efficacy of a universal smoking cessation intervention initiated in inpatient psychiatry and continued post-discharge: A randomised controlled trial. Australian & New Zealand Journal of Psychiatry, 2017. 51(4): p. 366–381.

38. Hertzog, M.A., Considerations in determining sample size for pilot studies. Research in nursing & health, 2008. 31(2): p. 180–191.

39. Senn, S., Testing for baseline balance in clinical trials. Statistics in medicine, 1994. 13(17): p. 1715–1726.

40. Braun, V. and V. Clarke, Thematic analysis. 2012: American Psychological Association.

