## Supplementary material for "Promoting Smoking Cessation and Preventing Relapse to Tobacco Use following a smokefree mental health in-patient stay (SCEPTRE feasibility study): a multi-centre randomised controlled feasibility study protocol": Fig 1 SCEPTRE CONSORT

### Slide 1
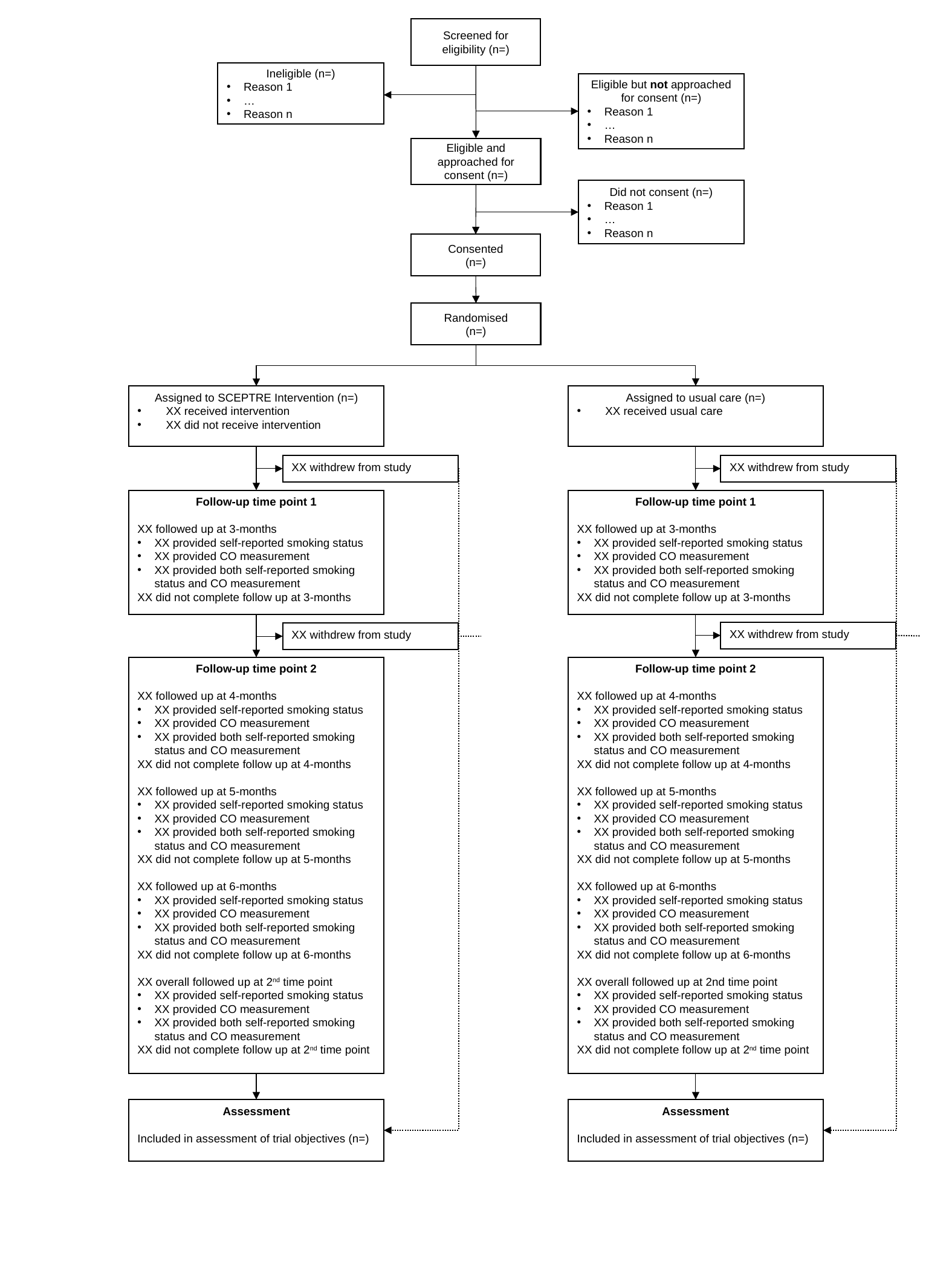

Screened for eligibility (n=)
Ineligible (n=)
Reason 1
…
Reason n
Eligible but not approached for consent (n=)
Reason 1
…
Reason n
Eligible and approached for consent (n=)
Did not consent (n=)
Reason 1
…
Reason n
Consented
(n=)
Randomised
(n=)
Assigned to usual care (n=)
XX received usual care
Assigned to SCEPTRE Intervention (n=)
XX received intervention
XX did not receive intervention
XX withdrew from study
XX withdrew from study
Follow-up time point 1
XX followed up at 3-months
XX provided self-reported smoking status
XX provided CO measurement
XX provided both self-reported smoking status and CO measurement
XX did not complete follow up at 3-months
Follow-up time point 1
XX followed up at 3-months
XX provided self-reported smoking status
XX provided CO measurement
XX provided both self-reported smoking status and CO measurement
XX did not complete follow up at 3-months
XX withdrew from study
XX withdrew from study
Follow-up time point 2
XX followed up at 4-months
XX provided self-reported smoking status
XX provided CO measurement
XX provided both self-reported smoking status and CO measurement
XX did not complete follow up at 4-months
XX followed up at 5-months
XX provided self-reported smoking status
XX provided CO measurement
XX provided both self-reported smoking status and CO measurement
XX did not complete follow up at 5-months
XX followed up at 6-months
XX provided self-reported smoking status
XX provided CO measurement
XX provided both self-reported smoking status and CO measurement
XX did not complete follow up at 6-months
XX overall followed up at 2^nd^ time point
XX provided self-reported smoking status
XX provided CO measurement
XX provided both self-reported smoking status and CO measurement
XX did not complete follow up at 2^nd^ time point
Follow-up time point 2
XX followed up at 4-months
XX provided self-reported smoking status
XX provided CO measurement
XX provided both self-reported smoking status and CO measurement
XX did not complete follow up at 4-months
XX followed up at 5-months
XX provided self-reported smoking status
XX provided CO measurement
XX provided both self-reported smoking status and CO measurement
XX did not complete follow up at 5-months
XX followed up at 6-months
XX provided self-reported smoking status
XX provided CO measurement
XX provided both self-reported smoking status and CO measurement
XX did not complete follow up at 6-months
XX overall followed up at 2nd time point
XX provided self-reported smoking status
XX provided CO measurement
XX provided both self-reported smoking status and CO measurement
XX did not complete follow up at 2^nd^ time point
Assessment
Included in assessment of trial objectives (n=)
Assessment
Included in assessment of trial objectives (n=)
