## Supplementary material for "Promoting Smoking Cessation and Preventing Relapse to Tobacco Use following a smokefree mental health in-patient stay (SCEPTRE feasibility study): a multi-centre randomised controlled feasibility study protocol": Fig 2 SCEPTRE intervention pathway

**My-Try Specialist**

Supervision and support

Training

**Inpatient (commencing pre-discharge)**

**Pre-discharge reflection and evaluation:** Exploring tobacco use and previous quit attempts, including biopsychosocial aspects relevant to discharge and CO measurement

**Pre-discharge assessment:** Goal setting and planning

**Smoke Free app onboarding session:** Participant-led training and familiarisation session on how to use the app if chosen at discharge

**On discharge from inpatient setting**

Provision of NRT or e-cigarettes for use at home (dependent on usual care)

Information on securing NRT/e-cigarette supply post-discharge

**Post-discharge**

Tailored behavioural support calls (telephone or video call)

Offer of use of the Smoke Free app

Motivational text messaging

**Figure 2.** SCEPTRE intervention pathway

Tailored resource folder (My-Try Kit)
