## Supplementary material for "Promoting Smoking Cessation and Preventing Relapse to Tobacco Use following a smokefree mental health in-patient stay (SCEPTRE feasibility study): a multi-centre randomised controlled feasibility study protocol": Fig 3 SPIRIT

Figure 3. Standard Protocol Items: Recommendations for Interventional Trials (SPIRIT) Figure – schedule of enrolment, intervention and data collection

|  | **STUDY PERIOD** | | | | |
| --- | --- | --- | --- | --- | --- |
|  | **Enrolment** | **Allocation** | **Post-allocation** | | **Close-out** |
| **TIMEPOINT**** | ***-t_1_*** | **0** | ***3 months*** | ***4/5/6 months*** | ***Post follow up*** |
| **ENROLMENT:** |  |  |  |  |  |
| **Eligibility screen** | X |  |  |  |  |
| **Informed consent** | X |  |  |  |  |
| **Allocation** |  | X |  |  |  |
| **INTERVENTIONS:** |  |  |  |  |  |
| ***[SCEPTRE*]*** |  |  |  |  |  |
| ***[Usual Care]*** |  |  |  |  |  |
| **ASSESSMENTS:** |  |  |  |  |  |
| ***Baseline variables*** |  |  |  |  |  |
| *Gender* |  | X |  |  |  |
| *Age* |  | X |  |  |  |
| *Marital Status* |  | X |  |  |  |
| *Ethnicity* |  | X |  |  |  |
| *Primary mental health diagnosis* |  | X |  |  |  |
| *Smoking status* |  | X |  |  |  |
| *Housing/accommodation status* |  | X |  |  |  |
| ***Proposed trial outcome measures*** |  |  |  |  |  |
| ***Smoking history and behaviour prior admission*** |  |  |  |  |  |
| *number of cigarettes smoked per day* |  | X | X | X |  |
| *number of past quit attempts* |  | X | X | X |  |
| *e-cigarettes and NRT* |  | X | X | X |  |
| ***Smoking history and behaviour during inpatient stay*** |  |  |  |  |  |
| *number of cigarettes smoked per day* |  | X | X | X |  |
| *greatest length of time abstinent during stay* |  | X | X | X |  |
| *HSI* |  | X | X | X |  |
| *Strength of urges to smoke* |  | X | X | X |  |
| *Motivation to Quit Questionnaire* |  | X | X | X |  |
| *e-cigarettes and NRT* |  | X | X | X |  |
| *(CO) reading* |  | X | X | X |  |
| ***Secondary outcome measures*** |  |  |  |  |  |
| *PHQ-9* |  | X | X | X |  |
| *GAD-7* |  | X | X | X |  |
| *EQ-5D-5L* |  | X | X | X |  |
| *Frequency of use of health services* |  | X | X | X |  |
| ***Acceptability measures*** |  |  |  |  |  |
| *Acceptability of study design and procedures* |  |  | X | X |  |
| *Qualitative interviews* |  |  |  |  | X |
| *MTS competency* |  |  |  |  |  |

* Promoting **S**moking **CE**ssation and **P**reven**T**ing **RE**lapse to Tobacco use following a smokefree mental health in-patient stay
