## Supplementary material for "Promoting Smoking Cessation and Preventing Relapse to Tobacco Use following a smokefree mental health in-patient stay (SCEPTRE feasibility study): a multi-centre randomised controlled feasibility study protocol": Supp 2 Consent Form

**SCEPTRE: Participant Consent Form**

| **Participant ID number:** |
| --- |

**Title of Project:** A randomised, controlled feasibility study of the SCEPTRE intervention to support smoking and prevent relapse to tobacco use following a smoke free mental health inpatient stay

If you wish to take part in the SCEPTRE study, **please initial each of the boxes below, sign and date this form.**

|  | | **Initial** |
| --- | --- | --- |
| **1** | I confirm that I have read and understand the information sheet version [*XX*] dated [*XX*] for the above study and have had the opportunity to ask any questions about the study and any questions have been answered to my satisfaction. |  |
| **2** | I understand that my participation is voluntary and that I am free to withdraw at any time without giving any reason, and without my medical care or legal rights being affected. |  |
| **3** | I agree to the University of York and the NHS trust holding copies of my consent form, other study related documents and my contact details to allow them to contact me to complete questionnaires and/or attend appointments. |  |
| **4** | I agree to my GP being informed of my participation in the study and being advised of any significant information relating to my health that comes to light during my participation. |  |
| **5** | I understand that relevant sections of my hospital medical notes and data collected during the study, may be looked at by individuals from the University of York, from regulatory authorities, the study Sponsor (Sheffield Health & Social Care NHS Foundation or from the NHS Trust, where it is relevant to my taking part in this research. I give permission for these individuals to have access to my records. |  |
| **6** | I understand that the study data may be stored and used in relevant future research, including by researchers in other institutions, but the data will not be used or released in such a way that I could be identified. |  |
| **7** | **I agree to take part in the SCEPTRE study.** |  |

**Optional consent**

In addition to the above statements please initial the following box(es) to indicate whether you agree with the following statement(s). Your participation in this research study will not be affected if you do not agree with these statements.

|  | | **Yes** | **No** |
| --- | --- | --- | --- |
| **8** | I agree to take part in an interview with a member of the research team to give my opinion about the SCEPTRE support package I have received. I agree for these to be audio recorded and transcribed. I understand that I can ask the member of the research team to stop the interview at any time without giving a reason. |  |  |
| **9** | I agree to be contacted about future relevant research studies and give permission for storage of my contact information for this purpose. |  |  |
| **10.** | I would like to be informed of the study results and give permission for storage of my contact information for this purpose. |  |  |

| …………………………………………………………. | ……………………………. | ……………………………………………… |
| --- | --- | --- |
| Name of participant | Date (dd/mm/yyyy) | Signature |

| …………………………………………………………. | ……………………………. | ……………………………………………… |
| --- | --- | --- |
| Name of person taking consent | Date (dd/mm/yyyy) | Signature |

If applicable

| …………………………………………………………. | ……………………………. | ……………………………………………… |
| --- | --- | --- |
| Name of witness | Date (dd/mm/yyyy) | Signature |

The original consent form should be kept in the site file and a copy to (tick): ☐ York Trials Unit ☐ Participant ☐ Clinical Notes
